# Prevalence of malnutrition and associated factors among adult patients on Antiretroviral Therapy follow up care in Jimma Medical Center, Southwest Ethiopia

**DOI:** 10.1101/19011130

**Authors:** Dawit Wolde Daka, Meskerem Seboka Ergiba

## Abstract

**Background:** Malnutrition especially under nutrition is the main problem that is seen over people living with HIV/AIDS and can occur at any age. Multiple factors contributed to malnutrition of HIV/AIDS patients and it need immediate identification and prompt action. The objective of this study was to assess the nutritional status of patients and identify factors associated with malnutrition among HIV/AIDS patients on follow-up care in Jimma medical center, Southwest Ethiopia.

**Methods:** A cross-sectional study design was conducted from March-April, 2016. Data was collected retrospectively from clinical records of HIV/AIDS patients enrolled for follow up care in ART clinic from June 2010 to January 2016. Binary and multiple variable logistic regression was done to identify independent predictor of malnutrition.

**Results:** Data of 971 patients were included in the study. The prevalence of under nutrition (BMI<18.5) was (36.8%) (95% CI: 33.8%-39.8%) and out of which severe malnutrition accounts 9.7%. Overweight and obese was 8.6%. Malnutrition was more likely among widowed patients (AOR=1.7, 95% CI, 1.034-2.798), patients in the WHO clinical AIDS staging of three (AOR=2.3, 95% CI, 1.392-3.693) and four (AOR=3.2, 95% CI, 1.667-5.943), patients with CD4 cell count of <200 cells/mm^3^ (AOR=2.0, 95% CI, 1.463-2.837) and patients with a functional status of bedridden (AOR=4.677, 95% CI, 1.761-12.419) and ambulatory (AOR=2.763, 95% CI, 1.833-4.165).

**Conclusion:** Both under nutrition and overweight are prevalent among HIV/AIDS patients in Jimma Medical Center, Ethiopia. Malnutrition was significantly associated with clinical outcome of patients. Hence, nutritional assessment, care and support should be strengthened. Critical identification of malnourished patients and prompt interventions should be undertaken.

## Introduction

Human immune deficiency virus (HIV) and malnutrition has multifaceted and multidirectional relationships. Both are related each other in causing progressive damage to the immune system. HIV compromises nutritional status and poor nutrition further weakness the immune system of individuals, increasing susceptibility to opportunistic infections. HIV can cause or worsen malnutrition by causing reduced food uptake, increased energy requirements and poor nutrient absorptions(1–3).

Poor nutritional status is one of the major complications of HIV and a significant factor in a full-blown AIDS. In many resource limited settings, many people who become infected with HIV may already be undernourished due to factors such as unemployment and difficulty to procure food. Food insecurity remains the major challenge in the progresses made to avert morbidity and mortality attributed to HIV/AIDS. Food insecurity and malnutrition increase high risk sexual behaviors, inconsistent condom use and multiple partnership. On the other hand, malnutrition leads to ART non-adherence among HIV/AIDS patients (4,5).

In Sub-Saharan Africa region, malnutrition (both under nutrition and overweight) is prevalent among HIV/AIDS patients enrolled in care (6–8). The prevalence of adult under nutrition is 19% in Tanzania(6), 10% in Zimbabwe(7) and 19% in Senegal(9). The prevalence of malnutrition is also high in Ethiopia; 25.2% in Butajira hospital(10) and 12.3% in Dilla university hospital(11). The main determinants of malnutrition ranges from individual level factors to underlying factors. Prevalence of malnutrition is determined by wealth index and educational attainment, where malnutrition decreased with increase in wealth index and educational attainment(12). A lower CD4 cell count, sex, age, advanced HIV diseases, presence of opportunistic infections, adherence concern, inability to access food and having social support were also significantly associated with malnutrition (6,7,9,13). Malnutrition is less likely in Females and older ages (35-44 years and ≥45 years) compared to 15-24 years. Having social support and informal care giving had also reduced the odds of under nutrition(13). On the contrary, malnutrition is more likely in those who had advanced HIV disease(6,7,9).

The high prevalence of HIV/AIDS(14) compounded by high rates of malnutrition remains the main challenge of health systems in SSA region. Currently, beyond under nutrition, overweight is also the main problem among HIV/AIDS patients. Despite improvement in treatment coverage over time(14), treatment effectiveness remains low due to factors such as non-adherence, quality and nutritional status of patients (15,16). The probability of mortality is highest among patients with poor nutritional status, as malnutrition is the cause for damage in the immune system and subsequent susceptibility to illnesses(17).

With encouraging effort from government and partners, Ethiopia has achieved encouraging results by reducing HIV/AIDS morbidity and mortality. Between 2000 and 2017, new HIV infections reduced by 90% and AIDS related mortality among adults reduced by more than 50%. While, HIV treatment coverage was reached to 71% in 2017(18). Though, the prevalence of HIV/AIDS remains high (0.9%) (19) and in 2017 there were an estimated 613,000 people living with HIV/AIDS, out of which 29% lacked access to treatments. Ethiopia remained one among the 25 countries with the highest number of new HIV infections worldwide. Out of the 25 countries, 17 were located in Africa region(18).

HIV infection and poor nutritional status are interlinked and strengthening nutritional assessment, care and support is essential to improve the effectiveness of HIV treatments. This requires generating evidences on the prevalence of malnutrition and factors affecting it. Hence, this study was aimed to estimate the prevalence of malnutrition (under nutrition) among adult HIV/AIDS patients enrolled in HIV/AIDS care in the period between 2010 and 2016 at Jimma Medical Center, a tertiary hospital in Ethiopia with an established HIV/AIDS treatment program.

## Materials and Methods

### Study setting

Jimma Medical Center, formerly known as Jimma University Specialized Hospital, was one of the teaching hospitals in Ethiopia located in Jimma city administration, 355km Southwest direction of the capital city, Addis Ababa. It is a government hospital which is an affiliate of Jimma University providing trainings for health science students in a range of disciples. The hospital also provides a higher level of clinical care for around 15 million catchment population located in Southwest part of Ethiopia. The hospital has 36 departments, of which ART clinic was one among them. Since 2005, the hospital has been providing highly active antiretroviral therapy (HAART) for people living with HIV/AIDS (PLWHA). During the study period (July 2010 to January 2016), about a total of 5554 patients were on HAART.

### Study design

A cross-sectional study design was conducted. Data was collected retrospectively from clinical records of HIV/AIDS patients enrolled for follow up care in ART clinic from July 2010 to January 2016. Data was retrieved from Pre-ART and ART log books of adults (>15 years). Data was extracted from March to April 2016.

### Study participants selection

The study participants were cohort of HIV/AIDS patients on follow-up care who are eligible for this study. The study participants were selected by using a systematic sampling strategy; where the clinical records of ART clients from July 2010 to January 2016 was used as a sampling frame. First, a sequential numbers starting from 1001 was provided to each records. The first record of patient was selected using lottery method and every K^th^ records was identified for data abstraction. HIV/AIDS patients who have a follow-up care on the same facility and that have key baseline information’s such as demographic characteristics (age, ethnicity, and marital status) and clinical information (such as CD4 count, viral load, WHO clinical stage, weight, height, and hemoglobin level) were eligible for this study.

The sample size was computed using OpenEpi Version 3 sample size calculator for proportions by using assumptions of a 95% CI, the outcome factor in the population of 46.8%(20) and confidence limit of 3%. The calculated sample size yields 1062.

### Measurement

A data abstraction tool, having the same structure with ART register and patient cards, was used to collect data. The tool comprised of components such as patient back ground information, immunologic status, clinical and laboratory examinations (CD4 count, viral load, WHO clinical stage, weight, height) and antiretroviral regimen and opportunistic infection prophylaxis. Three clinicians who had also trained on ART service provision were recruited for data collection. Another clinician also supervised the overall data collection processes.

### Statistical analysis

Data was cleaned for inconsistency and incompleteness. Then it was entered to epidata version 3.1 and imported to SPSS version 21. Descriptive statistics was conducted and variables were presented using mean, frequencies and proportion. Body Mass Index (BMI) was calculated as weight in kilograms divided by the squares of height in meters (kg/m^2^). For the initial analysis, BMI was stratified into the WHO criteria: <18.5 (under nutrition), >18.5 to 25 (normal nutrition) and >25 kg/m^2^ (overweight and obese). In this study under nutrition or BMI (<18.5 kg/m^2^) was estimated (3). Independent predictors of adult malnutrition, such as socio-demographic and clinical characteristics, were determined using binary and multiple variable logistic regression. Covariates for the multiple variable analysis were selected using Backward LR method. While, the model fitness (goodness) was assessed by using Wald test for individual variables and maximum likelihood ratio test for overall model.

### Ethical consideration

Research protocol was submitted to Institute of Health of Jimma university and get ethical approval (Ref no. RPGC/26/2016; March, 2016). Research permission was obtained from Jimma Medical Center. Confidentiality of patient information was maintained though use of codes, where name and personal identifiers of patients was not recorded. Paper based data was kept in a locked cabinet and computer based data was secured with passwords. Except the research team member, any one couldn’t access patient data.

## Results

### Participation

A total of 971 complete ART registrations and client records which incorporated basic information’s were included in the analysis, making a document completion rate of 91%.

### Socio-demographic characteristics

Table 1 shows the socio-demographic characteristics of patients on HIV/AIDS follow care. Most of the study participants were Female (60%) and in the age group of <30 years (41%). Nearly half (48.2%) of them were married and over half were followers of orthodox religion (58%). While, 89% were unemployed and slightly over 4 in 10 (43.5%) of them had attended primary education (**Table 1**).

**Table 1.** Socio-demographic characteristics of patients on HIV/AIDS follow up care at Jimma Medical Center, Southwest Ethiopia, 2016.

| **Variables** | **Frequency** | **%** |
| --- | --- | --- |
| Gender |  |  |
| **Male** | 372 | 40 |
| **Female** | 558 | 60 |
| **Total** | 930 | 100 |
| Age (in years) |  |  |
| **<30** | 402 | 41.4 |
| **30-39** | 368 | 37.9 |
| **40-49** | 151 | 15.6 |
| **>=50** | 50 | 5.1 |
| **Total** | 971 | 100 |
| Marital status |  |  |
| **Married** | 464 | 48.2 |
| **Never married** | 164 | 17.0 |
| **Separated** | 136 | 14.1 |
| **Divorced** | 82 | 8.5 |
| **Widowed** | 116 | 12.1 |
| **Total** | 962 | 100 |
| Religion |  |  |
| **Muslim** | 302 | 31.9 |
| **Orthodox** | 549 | 58.0 |
| **Protestant** | 90 | 9.5 |
| **Catholic** | 6 | 0.6 |
| **Total** | 947 | 100 |
| Educational status |  |  |
| **No education** | 188 | 19.5 |
| **Primary education** | 418 | 43.5 |
| **Secondary education** | 266 | 27.7 |
| **Tertiary education** | 90 | 9.4 |
| **Total** | 962 | 100 |
| Employment status |  |  |
| **Employed** | 95 | 10.5 |
| **Unemployed** | 814 | 89.5 |
| **Total** | 909 | 100 |
| Running water |  |  |
| **Available** | 771 | 83.4 |
| **Not available** | 154 | 16.6 |
| **Total** | 925 | 100 |
| Electricity |  |  |
| **Available** | 901 | 97.2 |
| **Not available** | 26 | 2.8 |
| **Total** | 927 | 100 |

### Clinical Characteristics

Table 2 shows the clinical characteristics, HIV/AIDS disclosure status, access to spiritual care giver and HIV support group, counseling and health education on HIV, and adherence concern of patients on HIV/AIDS follow care. Majority of patients disclosed their status (87%) and slightly over 3/4^th^ of them had attended HIV related health education sessions (76.5%). Nearly a third (29%) of them were in the WHO clinical stage of II and another 33% were at WHO stage of III at start of ART. Moreover, slightly over half (55.8%) have CD4 count <200 and over 3/4^th^ (77.6%) of them had a working functional status at start of ART (**Table 2**).

**Table 2.** Clinical and service related characteristics of patients on ART follow up care in Jimma Medical Center, 2016.

| **Variables** | **Frequency** | **Percent** |
| --- | --- | --- |
| Disclosure status |  |  |
| **Yes** | 796 | 87.0 |
| **No** | 119 | 13.0 |
| **Total** | 915 | 100 |
| Access to spiritual caregiver |  |  |
| **Yes** | 490 | 53.1 |
| **No** | 432 | 46.9 |
| **Total** | 922 | 100 |
| Access to community support/HIV support group |  |  |
| **Yes** | 275 | 29.9 |
| **No** | 646 | 70.1 |
| **Total** | 921 | 100 |
| Attended HIV related counseling session |  |  |
| **Yes** | 389 | 42.3 |
| **No** | 531 | 57.7 |
| **Total** | 920 | 100 |
| Attended HIV related HE session |  |  |
| **Yes** | 705 | 76.5 |
| **No** | 217 | 23.5 |
| **Total** | 922 | 100 |
| Adherence concern |  |  |
| **Stigma** | 584 | 70.9 |
| **Afraid of medications(side effects)** | 94 | 11.4 |
| **Doubt that medications couldn't work** | 51 | 6.2 |
| **Depression or anxious** | 68 | 8.3 |
| **Forget to take medication and other reasons** | 27 | 3.2 |
| **Total** | 824 | 100 |
| WHO Clinical AIDS staging |  |  |
| **Stage one** | 232 | 24.4 |
| **Stage two** | 283 | 29.7 |
| **Stage three** | 320 | 33.6 |
| **Stage four** | 117 | 12.3 |
| **Total** | 952 | 100 |
| CD4 cell count |  |  |
| **<200** | 506 | 55.8 |
| **>=200** | 400 | 44.2 |
| **Total** | 906 | 100 |
| Diagnosed for active TB |  |  |
| **Yes** | 124 | 13.2 |
| **No** | 817 | 86.8 |
| **Total** | 941 | 100 |
| Functional status at start of ART |  |  |
| **Ambulatory** | 175 | 19.1 |
| **Bedridden** | 30 | 3.3 |
| **Working** | 711 | 77.6 |
| **Total** | 916 | 100 |
| Patient past opportunistic infection |  |  |
| **Yes** | 805 | 82.9 |
| **No** | 166 | 17.1 |
| **Total** | 971 | 100 |

### Prevalence of Malnutrition

Table 3 shows the prevalence of malnutrition among the socio-demographic and clinical characteristics of patients. The prevalence of malnutrition (under nutrition) in the study area was 357(36.8%) (95% CI: 33.8%-39.8%). Out of which, severe malnutrition accounts 94(9.7%) and moderate malnutrition accounts of 263(27.1%). While, the prevalence of overweight and obese was 8.6%.

**Table 3.** Socio-demographic and clinical characteristics of study participants grouped by nutritional status (BMI^a^) in Jimma Medical Center, Southwest Ethiopia, 2016. **Legend**. BMI^a^ = Body Mass Index

| **Variable** | **Normal nutritional status**<br>**No (%)** | **Under nutrition**<br>**No (%)** |
| --- | --- | --- |
| Gender |  |  |
| **Male** | 225(38.5) | 147(42.5) |
| **Female** | 359(61.5) | 199(57.5) |
| **Total** | 584(100) | 346(100) |
| Age group (years) |  |  |
| **<30** | 253(41.2) | 149(41.7) |
| **30-39** | 227(37) | 141(39.5) |
| **40-49** | 100(16.3) | 51(14.3) |
| **>=50** | 34(5.5) | 16(4.5) |
| **Total** | 614(100) | 357(100) |
| Marital status |  |  |
| **Married** | 308(50.6) | 156(44.2) |
| **Never married/ single** | 98(16.1) | 66(18.7) |
| **Separated** | 88(14.4) | 48(13.6) |
| **Divorced** | 52(8.5) | 30(8.5) |
| **Widowed** | 63(10.4) | 53(15) |
| **Total** | 609(100) | 353(100) |
| Religious status |  |  |
| **Muslim** | 185(30.9) | 117(33.6) |
| **Orthodox** | 359(59.9) | 190(54.6) |
| **Protestant** | 51(8.5) | 39(11.2) |
| **Catholic** | 4(0.7) | 2(0.6) |
| **Total** | 599(100) | 348(100) |
| Educational status |  |  |
| **No education** | 126(20.7) | 62(17.6) |
| **Primary education** | 263(43.1) | 155(44.0) |
| **Secondary education** | 164(26.9) | 102(29.0) |
| **Tertiary education** | 57(9.3) | 33(9.0) |
| **Total** | 610(100) | 352(100) |
| Employment status |  |  |
| **Employed** | 64(11.1) | 31(9.3) |
| **Unemployed** | 510(88.9) | 304(90.7) |
| **Total** | 574(100) | 335(100) |
| WHO Clinical AIDS staging |  |  |
| **Stage one** | 174(29) | 58(16.5) |
| **Stage two** | 212(35.3) | 71(20.2) |
| **Stage three** | 167(27.8) | 153(43.5) |
| **Stage four** | 47(7.8) | 70(19.9) |
| **Total** | 600(100) | 352(100) |
| CD4 cell count |  |  |
| **<200** | 277(48.4) | 229(68.6) |
| **≥200** | 295(51.6) | 105(31.4) |
| **Total** | 572(100) | 334(100) |
| Functional status |  |  |
| **Ambulatory** | 68(11.7) | 107(31.8) |
| **Bed ridden** | 9(1.6) | 21(6.3) |
| **Working** | 503(86.7) | 208(61.9) |
| **Total** | 580(100) | 336(100) |
| Patient past opportunistic infection |  |  |
| **Yes** | 508(82.7) | 297(83.2) |
| **No** | 106(17.3) | 60(16.8) |
| **Total** | 614(100) | 357(100) |
| HIV/AIDS Disclosure status |  |  |
| **Yes** | 513(89.4) | 283(83) |
| **No** | 61(10.6) | 58(17) |
| **Total** | 574(100) | 341(100) |
| Access to spiritual care giver |  |  |
| **Yes** | 313(53.1) | 177(53.2) |
| **No** | 276(46.9) | 156(46.8) |
| **Total** | 589(100) | 333(100) |
| Access to community support group |  |  |
| **Yes** | 174(29.9) | 101(29.8) |
| **No** | 408(70.1) | 238(70.2) |
| **Total** | 582(100) | 339(100) |
| Attended HIV related counseling session |  |  |
| **Yes** | 244(42) | 145(42.8) |
| **No** | 337(58) | 194(57.2) |
| **Total** | 581(100) | 339(100) |
| Attended HIV related HE session |  |  |
| **Yes** | 459(78.9) | 246(72.4) |
| **No** | 123(21.1) | 94(27.6) |
| **Total** | 582(100) | 340(100) |
| Diagnosed for active TB |  |  |
| **Yes** | 63(10.6) | 61(17.6) |
| **No** | 531(89.4) | 286(82.4) |
| **Total** | 594(100) | 347(100) |

Under nutrition was more prevalent in female (57.5%), <30 years age group (41.7%), married (44.2%), orthodox religious follower (54.6%), primary education status (44%) and unemployed patients (90.4%) (**Table 3**).

### Socio-demographic and clinical factors associated with malnutrition

Table 4 and 5 shows binary and multiple variable analysis of different independent variables associated with malnutrition, respectively. In the binary logistic regression, only marital status was identified as a socio-demographic candidate variable for multiple variable logistic regression analysis. The remaining variables didn’t show any statistically significant association. The remaining were clinical variables including WHO clinical AIDS staging, CD4 cell count, functional status, diagnosed for active TB and patient attended HIV related health education sessions previously. In the multiple variable logistic regression, marital status, WHO clinical AIDS staging, CD4 cell count and patient functional status were identified as independent predictors of malnutrition.

**Table 4.** Bivariate association of different variables with malnutrition in PLWHA^b^ in Jimma Medical Center, Southwest Ethiopia, 2016. **Legend**. PLWHA^b^= People Living with HIV/AIDS, ^*^ P-value<0.05, COR^c^=Crude Odds Ratio, CI^d^= Confidence Interval

| Variables | Normal nutritional status<br>No (%) | Under nutrition<br>No (%) | COR<sup>c</sup> (95% CI<sup>d</sup>) |
| --- | --- | --- | --- |
| Gender |  |  |  |
| **Male** | 225(60.5) | 147(39.5) | 1 |
| **Female** | 359(64.3) | 199(35.7) | 0.848(0.647-1.112) |
| **Total** | 608(65.4) | 398(34.6) |  |
| Age group (years) |  |  |  |
| **<30** | 253(62.9) | 149(37.1) | 1 |
| **30-39** | 227(61.7) | 141(38.3) | 1.055(0.788-1.412) |
| **40-49** | 100(66.2) | 51(33.8) | 0.866(0.584-1.283) |
| **>=50** | 34(68) | 16(32) | 0.799(0.427-1.497) |
| **Total** | 614(63.2) | 357(36.8) |  |
| Marital status |  |  |  |
| **Married** | 308(66.4) | 156(33.6) | 1 |
| **Never married/ single** | 98(59.8) | 66(40.2) | 1.330(0.921-1.919) |
| **Separated** | 88(64.7) | 48(35.3) | 1.077(0.721-1.608) |
| **Divorced** | 52(63.4) | 30(36.6) | 1.139(0.699-1.857) |
| **Widowed** | 63(54.3) | 53(45.7) | 1.661(1.099-2.510)* |
| **Total** | 609(63.3) | 353(36.7) |  |
| Educational status |  |  |  |
| **No education** | 126(67) | 62(33) | 1 |
| **Primary education** | 263(62.9) | 155(37.1) | 1.198(0.833-1.722) |
| **Secondary education** | 164(61.7) | 102(38.3) | 1.264(0.854-1.870) |
| **Tertiary education** | 57(63.3) | 33(36.7) | 1.177(0.696-1.990) |
| **Total** | 610(63.4) | 352(36.6) |  |
| Employment status |  |  |  |
| **Employed** | 64(67.4) | 31(32.6) | 1 |
| **Unemployed** | 510(62.7) | 304(37.3) | 1.231(0.783-1.933) |
| **Total** | 574(63.1) | 335(36.9) |  |
| WHO Clinical AIDS staging |  |  |  |
| **Stage one** | 174(75) | 58(25) | 1 |
| **Stage two** | 212(74.9) | 71(25.1) | 1.005(0.673-1.5) |
| **Stage three** | 167(52.2) | 153(47.8) | 2.749(1.9-3.977)* |
| **Stage four** | 47(40.2) | 70(59.8) | 4.468(2.781-7.179)* |
| **Total** | 600(63) | 352(37) |  |
| CD4 cell count |  |  |  |
| **<200** | 277(54.7) | 229(45.3) | 2.323(1.750-3.083)* |
| **≥200** | 295(73.8) | 105(26.3) | 1 |
| **Total** | 572(63.1) | 334(36.9) |  |
| Functional status |  |  |  |
| **Ambulatory** | 68(38.9) | 107(61.1) | 3.805(2.697-5.369)* |
| **Bed ridden** | 9(30) | 21(70) | 5.643(2.542-12.525)* |
| **Working** | 503(70.7) | 208(29.3) | 1 |
| **Total** | 580(63.3) | 336(36.7) |  |
| Patient past opportunistic infection |  |  |  |
| **Yes** | 508(63.1) | 297(36.9) | 1.033(0.730-1.462) |
| **No** | 106(63.9) | 60(36.1) | 1 |
| **Total** | 614(63.2) | 357(36.8) |  |
| HIV/AIDS disclosure status |  |  |  |
| **Yes** | 513(64.4) | 283(35.6) | 0.580(0.394-0.855)* |
| **No** | 61(51.3) | 58(48.7) | 1 |
| **Total** | 574(62.7) | 341(37.3) |  |
| Access to spiritual care giver |  |  |  |
| **Yes** | 313(63.9) | 177(36.1) | 1.0(0.764-1.310) |
| **No** | 276(63.9) | 156(36.1) | 1 |
| **Total** | 589(63.9) | 333(36.1) |  |
| Access to community support group |  |  |  |
| **Yes** | 174(63.3) | 101(36.7) | 0.995(0.743-1.333) |
| **No** | 408(63.2) | 238(36.8) | 1 |
| **Total** | 582(63.2) | 339(36.8) |  |
| Attended HIV related counseling session |  |  |  |
| **Yes** | 244(62.7) | 145(37.3) | 1.032(0.787-1.354) |
| **No** | 337(63.5) | 194(36.5) | 1 |
| **Total** | 581(63.2) | 339(36.8) |  |
| Attended HIV related health education session |  |  |  |
| **Yes** | 459(65.1) | 246(34.9) | 0.701(0.514-0.956)* |
| **No** | 123(56.7) | 94(43.3) | 1 |
| **Total** | 582(63.1) | 340(36.9) |  |
| Diagnosed for active TB |  |  |  |
| **Yes** | 63(50.8) | 61(49.2) | 1.798(1.229-2.630)* |
| **No** | 531(65) | 286(35) | 1 |
| **Total** | 594(63.1) | 347(36.9) |  |

**Table 5.** Adjusted association of different variables with malnutrition in PLWHA^b^ in Jimma Medical Center, Southwest Ethiopia, 2016. **Legend**. ^b^ PLWHA=People Living with HIV/AIDS, AOR^e^=Adjusted Odds Ratio, CI^d^= Confidence Interval, ^*^ P-value<0.05

| Variables | Normal nutritional status<br>No (%) | Under nutrition<br>No (%) | AOR<sup>e</sup> for all variables (95%<br>CI<sup>d</sup>) |
| --- | --- | --- | --- |
| Marital status |  |  |  |
| **Married** | 308(66.4) | 156(33.6) | 1 |
| **Never married/ single** | 98(59.8) | 66(40.2) | 1.410(0.904-2.198) |
| **Separated** | 88(64.7) | 48(35.3) | 0.723(0.440-1.187) |
| **Divorced** | 52(63.4) | 30(36.6) | 1.506(0.835-2.716) |
| **Widowed** | 63(54.3) | 53(45.7) | 1.701(1.034-2.798)* |
| **Total** | 609(63.3) | 353(36.7) |  |
| WHO Clinical AIDS staging |  |  |  |
| **Stage one** | 174(75) | 58(25) | 1 |
| **Stage two** | 212(74.9) | 71(25.1) | 1.129(0.692-1.841) |
| **Stage three** | 167(52.2) | 153(47.8) | 2.267(1.392-3.693)* |
| **Stage four** | 47(40.2) | 70(59.8) | 3.147(1.667-5.943)* |
| **Total** | 600(63) | 352(37) |  |
| CD4 cell count |  |  |  |
| **<200** | 277(54.7) | 229(45.3) | 2.037(1.463-2.837)* |
| **≥200** | 295(73.8) | 105(26.3) | 1 |
| **Total** | 572(63.1) | 334(36.9) |  |
| Functional status |  |  |  |
| **Ambulatory** | 68(38.9) | 107(61.1) | 2.763(1.833-4.165)* |
| **Bed ridden** | 9(30) | 21(70) | 4.677(1.761-12.419)* |
| **Working** | 503(70.7) | 208(29.3) | 1 |
| **Total** | 580(63.3) | 336(36.7) |  |
| HIV/AIDS disclosure status |  |  |  |
| **Yes** | 513(64.4) | 283(35.6) | 0.847(0.505-1.421) |
| **No** | 61(51.3) | 58(48.7) | 1 |
| **Total** | 574(62.7) | 341(37.3) |  |
| Attended HIV related Health Education session |  |  |  |
| **Yes** | 459(65.1) | 246(34.9) | 0.715(0.495-1.033) |
| **No** | 123(56.7) | 94(43.3) | 1 |
| **Total** | 582(63.1) | 340(36.9) |  |
| Diagnosed for active TB |  |  |  |
| **Yes** | 63(50.8) | 61(49.2) | 0.874(0.537-1.423) |
| **No** | 531(65) | 286(35) | 1 |
| **Total** | 594(63.1) | 347(36.9) |  |

With regard to marital status, a greater number of malnourished patients were found in the group of the widowed (45.7%) followed by the single or never married ones (40.2%). Being a widow was significantly associated with malnutrition (AOR=1.7, 95% CI, 1.034-2.798).

HIV/AIDS patients in the WHO clinical AIDS staging three (47.8%) and four (59.8%) were more malnourished compared to patients in stage one and two. Being in the WHO clinical AIDS stage of three (AOR=2.3, 95% CI, 1.392-3.693) and four (AOR=3.2, 95% CI, 1.667-5.943) were significantly associated with malnutrition among adult HIV/AIDS patients in the study area.

Likewise, a greater number of malnourished patients had a CD4 cell count of <200cells/μL (45% versus 26%) and were bedridden (70%) and ambulatory (61%) in their functional status. There is a statistical significant association between patients CD4 cell count (AOR=2.0, 95% CI, 1.463-2.837) and their functional status with malnutrition. Being bedridden (AOR=4.677, 95% CI, 1.761-12.419) and ambulatory (AOR=2.763, 95% CI, 1.833-4.165) were significantly associated with malnutrition among adult HIV/AIDS patients in the study area.

In the binary logistic regression analysis, HIV AIDS disclosure status, being diagnosed for active TB and attended HIV related health education sessions were significantly associated with malnutrition. However, the association of these variables was not maintained after adjusting for all independent variables (**Table 4 and 5**).

## Discussion

This study was intended to estimate the prevalence of malnutrition and its determinants among adult HIV/AIDS patients who were enrolled on care or who were on ART treatments. The study had indicated that the overall prevalence of malnutrition (under nutrition) in the study area was 36.8% and out of which, severe malnutrition (<16 kg/m^2^) accounts of 9.7%. We found that, patient marital status, WHO clinical AIDS staging, CD4 cell count and patient functional status were significantly associated with malnutrition.

The prevalence of malnutrition was higher than a study conducted in different parts of Ethiopia; 12.3% in Dilla university hospital (11), 25.2% in Butajira hospital (10), 18.2% in Arba Minch area public health facilities (21), 30% in East Hararghe zone hospitals(22) and 27% in Nekemte referral hospital (23). Likewise, the prevalence is also much higher compared to studies done in different parts of the world; 19.5% in Tanzania(6), 10% in Zimbabwe(7) and 19.2% in Senegal(9). The difference in prevalence of malnutrition might be due to difference in socio-economic and other factors that may predispose the community to problem, such as food habit and culture. However, the prevalence of severe malnutrition (BMI<16kg/m^2^) was comparable to studies done at Tanzania (9%)(6) and Butajira hospital(9%)(10).

In our study under nutrition was more prevalent among female patients than male. This is comparable with other studies done in different parts of Ethiopia; Southern (11)and Eastern Ethiopia(22). Yet different from studies done in SSA countries (6,7) and slightly higher among male patients in study done in Butajira hospital, Southern Ethiopia(10). This implies that under nutrition is more common among females and this might be related to the socio-economic status of women, access to information and other predisposing factors that may affect food intake of women in the community. On the other hand, most women are already undernourished than men in the general population. In Ethiopia, only 51% of female attended school compared to 65% of men. Employment status was much higher among men than women. Likewise, most women were not accessed to mass media and internet compared to men. In Ethiopia, 24% of women aged 15-49 years were anemic and 22% were thin (BMI<18.5 kg/m^2^) (19).

Our study also indicated that, malnutrition is more prevalent in younger, married, primary education level and unemployed patients compared to their counter parts. As the age of patient’s increased, the prevalence of malnutrition was decreased in the study area. This is comparable with studies done in different parts of the world (6,7) and Ethiopia(10,22), where under nutrition is more prevalent among the younger adults and overweight is more prevalent among older patients. Additionally, studies also supports the higher prevalence of malnutrition among unemployed patients and patients in the lower educational status(11,24,25). While studies done in Northern and Southern Ethiopia indicated contradicting finding in which under nutrition was associated with greater age(11,26). Malnutrition was also much prevalent in patients on WHO clinical stage three and four, CD4 cell count less than 200 and in patients with a past opportunistic infections. This indicates interlink between HIV infection and poor nutritional status. HIV compromises the nutritional status and poor nutrition further weakens the immune system of individuals, increasing susceptibility to opportunistic infections(2).

In the multivariable logistic regression, patient marital status, WHO clinical AIDS staging, CD4 cell count and functional status were statistically associated with under nutrition in the study area. In the binary logistic regression variables such as diagnosis of active TB, attending HIV related health education sessions and disclosing HIV status were significantly associated with malnutrition. Though, these variables didn’t show a statistical significant associations while adjusted for other variables.

Our study revealed that, being a widow was significantly associated with malnutrition. HIV/AIDS patients who were in the WHO clinical stage of three and four were two times and three times more likely malnourished than those in the WHO clinical stage of one, respectively. In addition, malnutrition was also more likely among patients with CD4 cell count of <200 and patients with functional status of bedridden and ambulatory. Malnutrition was nearly five times and three times more likely among bedridden and ambulatory patients compared to patients who have a working functional status, respectively. Our finding was supported by other studies done in other areas else(6,7,10,11,13,21,22,24–26).

Malnutrition is the major factor in ensuring treatment effectiveness and thus, nutritional assessments, care and support for HIV/AIDS patients should be strengthened. Poor adherence is caused by lack of access to food or food insecurity. On the other hand, poor treatment adherence leads to infections and suppressed immunity of patients(9). Mortality is more likely among immune compromised HIV/AIDS patients and supplementation of therapeutic food can improve the nutritional status of patients(17).

Our study had assessed the prevalence of malnutrition on patients enrolled in care in the five successive periods (2010 and 2016). Moreover, adequate sample size was used to estimate malnutrition prevalence. Yet, our study has the following limitations. The study was based on secondary data (chart review) and thus may not address all of the variables that may affect malnutrition and may be subject to incomplete data bias.

## Conclusions

This finding indicated that the prevalence of malnutrition (both under nutrition and overweight) was high compared to other settings in Ethiopia. It was also indicated that WHO clinical AIDS staging three and four, CD4 cell count <200, functional status and marital status were significantly associated with malnutrition among adult HIV/AIDS patients. Though not statistically significant when adjusted to other variables, HIV disclosure status and access to HIV related health education were also associated with malnutrition. Hence, HIV treatment services should be supported with nutritional assessment, supplementation, counseling, care and support to patients. A comprehensive nutritional assessment and support should be provided for all patients on follow-up care. Moreover, community support to patients should be strengthened, as social determinants of health may also interact with effectiveness of treatments.

## Data Availability

Data is accessible and can be availed at any time on request. Please contact: to access data related to the manuscript. In addition, authors would like if medRxiv provides help in uploading data.

## Acknowledgments

The authors would like to thank Jimma Medical Center and Workers at Antiretroviral Therapy (ART) unit for cooperating with and supporting the research work.

## Authors Contributions

Conceived and designed the study: WD SM. Conducted the study: WD SM. Analyzed the data: WD. Wrote the paper: WD SM. Edited the manuscript: WD SM.

